# When strong mitigation against a pandemic backfires

**DOI:** 10.1101/2020.05.13.20100628

**Authors:** Pierre Chaigneau

## Abstract

I introduce social feasibility constraints in a SIR epidemiological model: at any point in time, the ability of a social planner to impose mitigation measures is limited, but it is increasing in the proportion of infected individuals. When considering threshold policies with constant levels of mitigation for a time period, the overall fatality rate in the population is non-monotonic in the levels of mitigation: higher levels of mitigation can increase the overall fatality rate. Intuitively, strong mitigation at a point in time can undermine the social feasibility of future mitigation.

> “When the economists talk the trade-off talk, lots of epidemiologists (and others) find it morally reprehensible when people are dying.”
>
> — Noah Feldman, Bloomberg, April 2 2020.

## 1 Introduction

Lockdowns have enormous economic costs. Yet their widespread adoption in 2020 has been justified on the basis that they will reduce contagion from the COVID-19 pandemic, as they did during the 1918 influenza pandemic (Hatchett, Mecher, and Lipsitch (2007)). The optimal lockdown policy^1^ can be derived in a SIR framework as the solution to an optimal control problem in which a social planner can at any point in time reduce new infections at a cost (Alvarez, Argente, and Lippi (2020), Piguillem and Shi (2020), Hall, Jones, and Klenow (2020), Hansen and Troy (2011),Jones, Philippon, and Venkateswaran (2020)). It is well-known that there is a tradeoff between the fatality rate in the population and the mitigation cost on the equilibrium path. On this equilibrium path, slightly more restrictive mitigation will reduce even further the fatality rate. This notion has driven advocacy for more restrictive lockdowns.

Yet, especially in an unexpected and unprecedented crisis, there is usually a gap between theoretically optimal policies and actual policies, Since the beginning of the COVID-19 pandemic, it has been obvious that many governments face obstacles when putting in place mitigation policies. This includes noncompliance among the population, especially at times when the perceived risk of infection is low, and implementation constraints which prevent continuous adjustments in the level of mitigation. Because of this gap between theoretically optimal and actual policies, properties of the former do not necessarily apply to the latter.

In this paper, I consider an infectious disease whose evolution can be described by the standard SIR epidemiological model. In this framework, I study the effect of various threshold mitigation policies in the presence of social feasibility constraints. The notion of social feasibility encompasses the notions of social acceptance and individual compliance. It varies across countries: many measures taken by China to suppress COVID-19 might not have been feasible in other countries including the US.^2^ In many countries, lockdowns have faced opposition and have been characterized with widespread noncompliance when the perceived risk of infection was low.^3^ Thus, effective mitigation measures can only be put in place under some conditions. To take this into account, I simply assume that there is an upper bound to the feasible level of mitigation which linearly depends on the proportion of infected individuals at a point in time: more infected individuals imply a higher risk of infection for susceptible individuals, which increases compliance and acceptance of mitigation measures, all else equal.

Within this framework, a hypothetical social planner can set the level of mitigation by decreasing the effective contact rate, i.e., the rate at which infected people transmit the disease, subject to social feasibility. To take into account the stepwise nature of actual mitigation policies, I consider threshold mitigation policies, which involve a constant mitigation level for a period of time. I consider “one-step”, “two-steps”, and “three-steps” threshold policies with one, two, and three levels of mitigation. These policies are consistent with “lockdowns” that were adopted by many countries in 2020 and followed by milder mitigation, and they can also generate “rolling lockdowns”.

The main result is that, for a wide range of social feasibility parameters, the overall fatality rate in the population is non-monotonic in the level(s) of mitigation. Very strong mitigation only minimizes the fatality rate when social feasibility is very high, and when it is followed in a timely manner by mild mitigation. Otherwise, reducing the effective contact rate beyond a certain level increases the overall fatality rate. Whereas a permanent reduction in the effective contact rate is unequivocally beneficial (“more infectious diseases are worse”), a temporary reduction in the effective contact rate that will reduce the number of fatalities in the short-term may still increase the total number of fatalities. Simply put, “erring on the side of caution” by setting a high level of mitigation can backfire. This has important implications for the design of mitigation policies and for the assessment of the policy response to COVID-19.

This surprising result can be explained as follows. With social feasibility constraints, mitigation measures must be lifted once the infection rate is sufficiently low. Strong mitigation measures bring down the infection rate rapidly without largely reducing the proportion of susceptible individuals. Unless strong mitigation is followed by well-calibrated mild mitigation, this reduction in the number of infected individuals puts an end to mitigation, which gets the effective reproduction number R of the disease above 1 and leads to a new spike in infections. This can result in “stop and go” mitigation, and in a higher overall fatality rate than lower-level mitigation. This emphasizes a so far overlooked cost of strong mitigation measures: by strongly reducing the infection rate, they can undermine the social feasibility of future mitigation and lead to a worse outcome overall.

In contrast to most of the epidemiology literature, which studies specific mitigation measures,I do not specify the nature of mitigation measures, and instead I only focus on the level of mitigation. There is a well-established literature on the prevention of influenza and pandemics. A recent influential paper by Ferguson et al. (2020) analyzes strategies against COVID-19, including “mitigation” which slows epidemic spread, and “suppression” which aims for very low levels of infections. They argue that suppression will require strong measures to be put in place until a vaccine is available, possibly on a “stop and go” basis.

This paper’s contribution to the burgeoning literature on the COVID-19 pandemic and to the epidemiological economics literature (Perrings et al. (2014)) is to take into account social feasibility and its consequences for the level and duration of mitigation. This perspective may help explain why the response to the COVID-19 pandemic and health outcomes varied across countries, and why a high level of mitigation may be effective in some social contexts but not in others.

## 2 Model

The framework relies on the standard SIRD model used in epidemiological research. At any time *t* ∈ [0,*T*], a population normalized at N =1 individuals is divided into those susceptible *S*(*t*), infected *I*(*t*), recovered *R*(*t*), and dead *D*(*t*), with *N* = *S*(*t*) + *I*(*t*) + *R*(*t*) + *D*(*t*) ∀*t*. The equations of motion for these components of the population are:

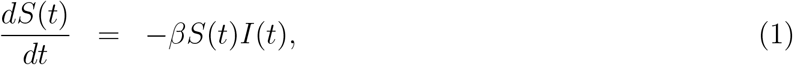

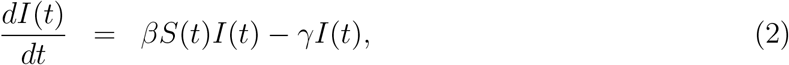

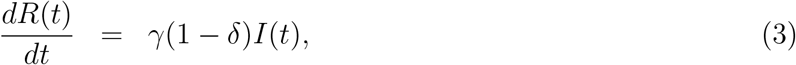

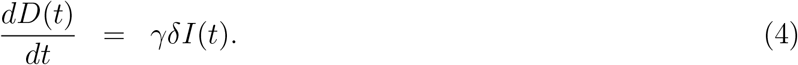

The three parameters are: the effective contact rate *β*; the removal rate *γ*; and the death rate for those infected *δ*.

We assume that *β* is not time invariant and can be altered from its baseline level *b* at a cost. Specifically, a social planner can take mitigation measures that reduce *β* by a factor *u*(*t*) ∈ [0,1] at time *t*, so that it is:

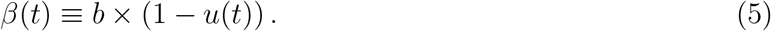

We assume that *R*(0) = *D*(0) = 0, so that the fatality rate in the population at time *T* is:

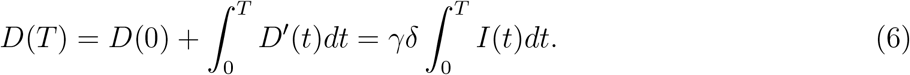

The model is calibrated for COVID-19. In the SIRD model, the basic reproduction number *R*_0_, which is the average number of additional infections generated by an infected individual, is equal to *β* times the duration of the infectious period. The CEBM at Oxford calculated from 16 studies a mean *R*_0_ of 2.65.^4^ The duration of the infectious period can be estimated to be 15 days on average, so that *γ* = 1/15 (Byrne et al. (2020)). Without any mitigation (u = 0), this gives *β* = *b* = *γR*_0_ ≈ 0.18. Based on the death toll and serological tests in New York city which indicated that 25 percent of the population had been infected with COVID-19, the fatality rate for infected individuals is set at 0.5%.^5^ The level of this parameter is not crucial for the main results (see section 4.1). Time is measured in days, and time 0 is when 0.1% of the population is infected.

## 3 Mitigation policies

### 3.1 Preliminary analysis

Figure 1 depicts the case with no mitigation, when the effective contact rate is: *β*(*t*) = *b* ∀*t*. Without any mitigation, the vast majority of the population gets infected, so that the death rate in the population, *D*(*T*) = 0.456%, is close to the death rate for infected people (*δ* = 0.5%).

**Figure 1:**
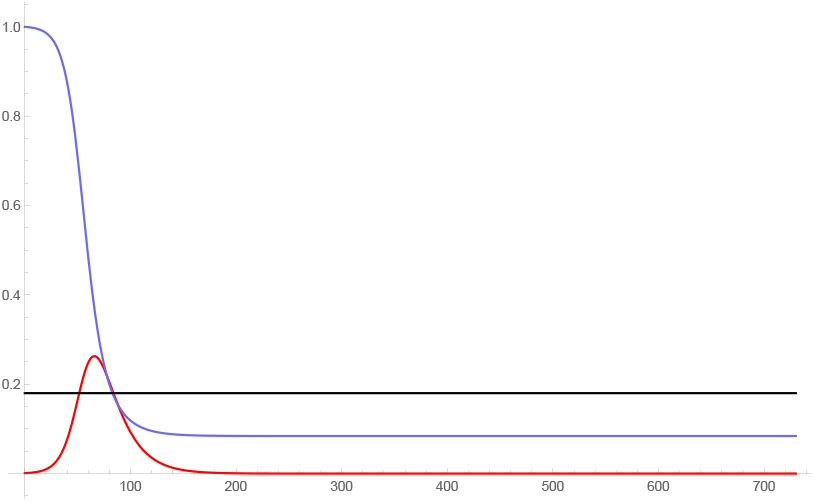
On the left, *I*(*t*) (red), *S*(*t*) (blue), and *b*(*t*) (black) for *u*(*t*) = 0 ∀*t* and *T* = 1, 000.

Figure 2 displays the case without social feasibility constraints when mitigation with constant level (*u*(*t*) = *u*_1_ ∀*t*) starts when 1% of the population is infected and only ends when 0% of the population is infected. In this case, unsurprisingly, the level of mitigation that minimizes the overall fatality rate is *u*_1_ = 100%.

**Figure 2:**
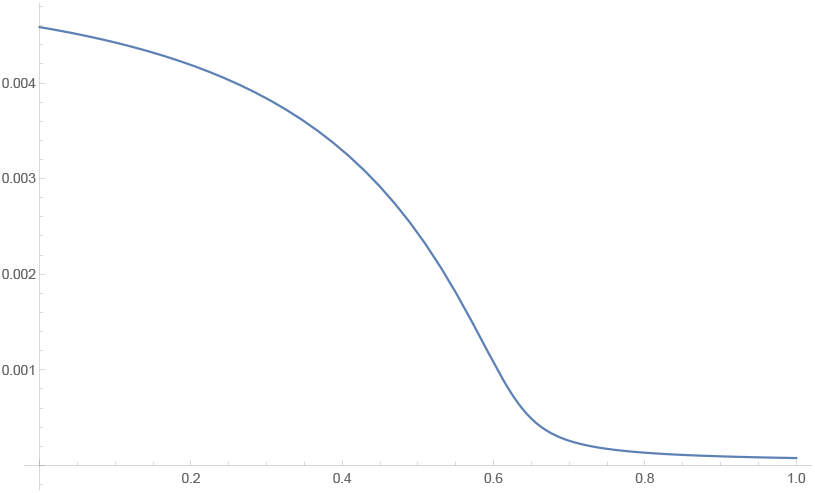
*D(T)* as a function of mitigation level *u*(*t*) = *u*_1_ for *u*_1_ ∈ [0, 1] and *T* = 1, 000 when mitigation starts when 1.0% of the population is infected and ends when 0.0% of the population is infected. The minimum is reached at *u*^*^_1_ = 100%, where *D*(*T*) = 0.008%.

### 3.2 Social feasibility

This section introduces social feasibility constraints.

China’s early suppression of COVID-19 relied on strong measures, which involved “forcibly isolating every resident in makeshift hospitals and temporary quarantine shelters.” According to Huiyao Wang, a senior adviser to China’s government, “You need to isolate people on an enormous scale.” Chinese officials “cast doubt on whether Americans could do what the Chinese did, for a mixture of reasons: political will and deep-rooted cultural inclinations among them.”^6^ According to Lawrence Gostin, a global health law scholar at Georgetown University, “China is unique in that it has a political system that can gain public compliance with extreme measures.”^7^

Mitigation measures taken against a pandemic are constrained by social acceptance and by the willingness of the population to comply. Indeed, “most measures for managing public health emergencies rely on public compliance for effectiveness.” (O’Malley, Rainford, and Thompson (2009)). Likewise, Besley and Velasco argue that “Because staying home and forgoing income or queuing two metres apart all have costs, people will follow lockdown and social distancing orders only if they view those orders, and the process that lead up to them,as legitimate.”^8^ In addition, recent evidence about COVID-19 briefly reviewed in a Supplementary Online Appendix suggests that the acceptance of strong measures as well as the willingness to comply at any point in time seems high enough only when the number of infections and deaths is high enough at this time. A large enough decrease in the level of infection in the population reduces the individual risk involved in violating or gaming lockdown orders as well as the social stigma attached to such actions. Consequently, public authorities face pressure to relax mitigation measures as well as enforcement issues once the proportion of infected individuals is sufficiently low.

To incorporate these considerations in a simple and transparent manner, I simply assume that the social planner faces two social feasibility constraints, which are parameterized by {*α_n_*,*α_c_*}. First, putting in place a new mitigation policy (switching from *u* = 0 to *u* > 0) requires a sufficiently high level of infection risk, and can only be achieved at time t when:

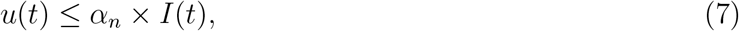

where *α_n_* ≥ 0. Second, when *u*(*t*) > 0, continuing a mitigation policy at time t requires the level of risk to be sufficiently high, but not as high as the level required to put in place a new policy:

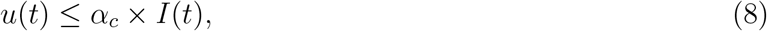

where *α_c_* > *α_n_* (see the Supplementary Online Appendix). The simple linear functional form in equations (7) and (8) allows social feasibility to be described by a small number of easily interpretable parameters. Both *α_n_* and *α_c_* are increasing in the acceptance and compliance with command-and-control instruments in the population, and in the perceived health risk involved in infection from a particular disease. *α_n_* = 0 means that no mitigation is ever possible (i.e. *u*(*t*) = 0 ∀*t*), whereas infinite values for *α_n_* and *α_c_* means that there is no social feasibility constraint.

### 3.3 One mitigation level

This section considers threshold mitigation policies with one mitigation level, i.e., u can take two values: 0 and *u*_1_ > 0, such that mitigation is in place whenever socially feasible. Mitigation starts (*u* = *u*_1_) when the proportion of infected individuals *I*(*t*) rises above the “mitigation threshold” corresponding to 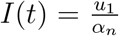 (see equation (7)), and ends (*u* = 0) when it falls below the “opening threshold” corresponding to 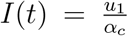 (see equation (8)). The social planner chooses the level of mitigation *u*_1_ ∈ [0, 1] to minimize the overall fatality rate in the population *D*(*T*). Note that the level of mitigation *u*_1_ chosen will affect the thresholds that determine when the mitigation period(s) start and end - the main results also hold when thresholds are exogeneously given, see the Supplementary Online Appendix.

**Figure 3:**
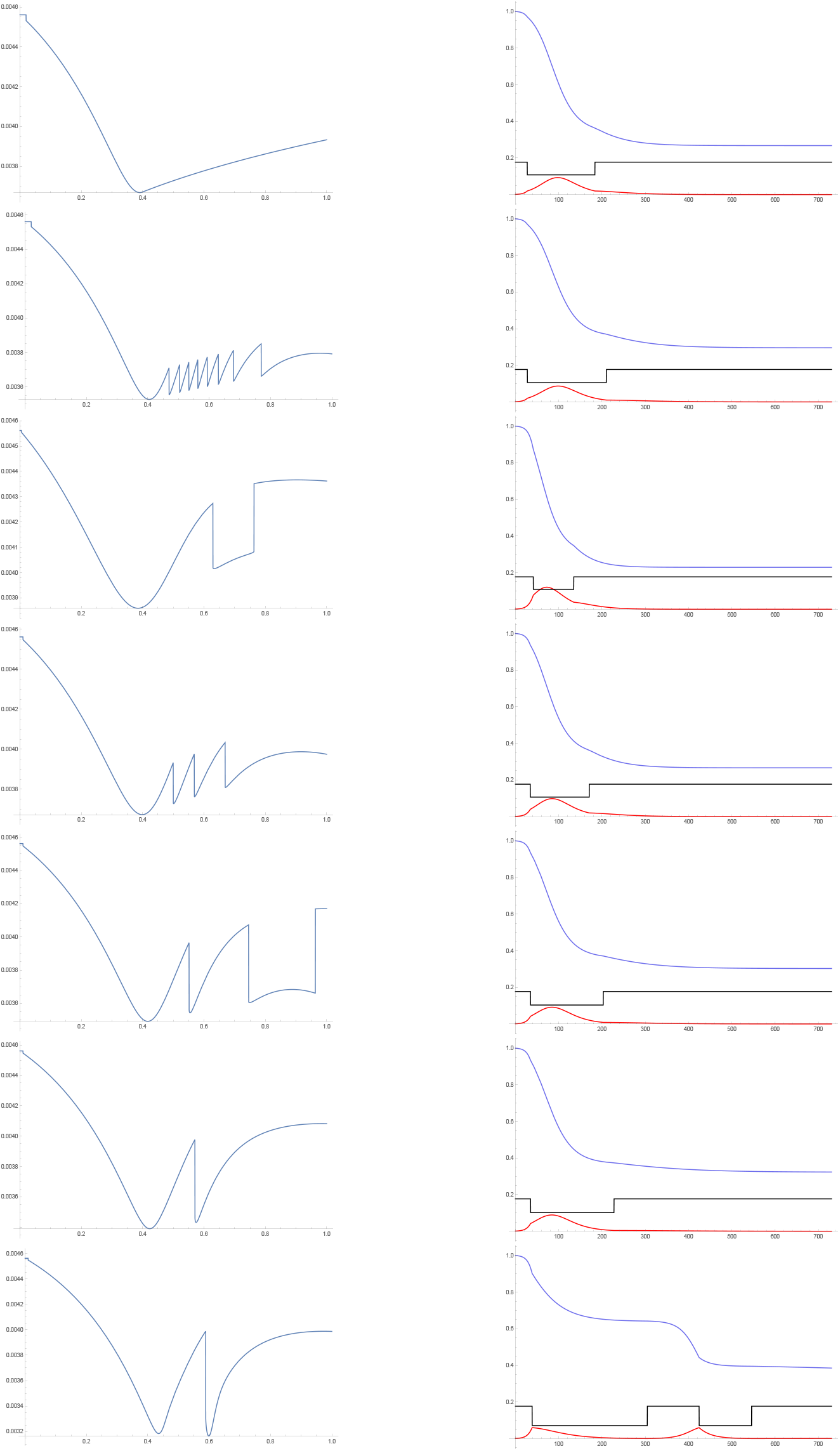
On the left, *D*(*T*) as a function of *u*_1_ for *u*_1_ ∈ [0, 1] and *T* = 1, 000. On the right, *I*(*t*) (red), *S*(*t*) (blue), and *b*(*t*) (black) for the level 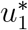 of *u_i_* that minimizes the overall fatality rate. On the first row, *α_n_* = 20, *α_c_* = 20.1, which gives 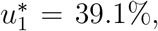, and *D*(*T*) = 0.367%. On the second row, *α_n_* = 20, *α_c_* = 40, which gives 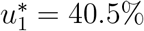, and *D*(*T*) = 0.353%. On the third row, *α_n_* = 5, *α_c_* = 10, which gives 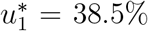, and *D*(*T*) = 0.386%. On the fourth row, *α_n_* = 10, *α_c_* = 20, which gives 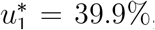, and *D*(*T*) = 0.367%. On the fifth row, *α_n_* = 10, *α_c_* = 50, which gives 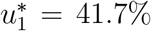, and *D*(*T*) = 0.349%. On the sixth row, *α_n_* = 10, *α_c_* = 100, which gives 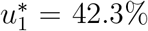, and *D*(*T*) = 0.339%. On the seventh row, *α_n_* = 10, *α_c_* = 1000, which gives 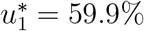, and *D*(*T*) = 0.316%.

A well-known lesson of the SIR model and its variants is that new infections at a given point in time will be less acute when either the number of infected individuals or the number of susceptible individuals is low. Without mitigation measures, the number of infected individuals rises sharply at a time when the number of susceptible individuals is still high, resulting in a very high rate of infections, and ultimately fatalities. Thus, “flattening the curve” is beneficial.

The main finding from the simulations described and depicted in Figure 3 is that, for a variety of social feasibility parameters {*α_n_*, *α_c_*}, setting the level of mitigation *u_1_* substantially higher than a cutoff around 40% does not substantially reduce the fatality rate. Unless mitigation is highly sustainable over time (*α_c_* is very high), setting *u*_1_ at a high level will result in a high fatality rate (see also the left panel in Figure 8 below). In the simulations depicted in Figure 3, the susceptibility curve decreases rather smoothly, and the infection rate remains low after the mitigation period ends, except in the last case.^9^ In the last case depicted in Figure 3, *α_c_* = 1000 allows for sustained mitigation. However, even then, at some point in time the opening threshold is still reached with strong mitigation 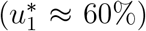. Since the proportion of susceptible individuals at this point is still high due to the preceding strong mitigation, this results in a spike in infections which triggers an additional mitigation period.

### 3.4 Two mitigation levels

This section considers policies with two mitigation levels, i.e., *u* can take three values: 0, *u*_1_ > 0, and *u*_2_ > 0, with 0 ≤ *u*_2_ < *u*_1_ ≤ 1. Having an additional level of mitigation u_2_ allows mitigation to continue at a lower level without reaching the opening threshold implicitly defined in equation (8). Accordingly, acute mitigation starts (*u* = *u*_1_) when the proportion of infected individuals rises above the “acute mitigation threshold” corresponding to 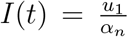(see equation (7)), mild mitigation starts (*u* = *u*_2_) when continuing with acute mitigation would violate social feasibility in equation (8), i.e., when *I*(*t*) falls below 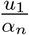 (the “mild mitigation threshold”), and mild mitigation ends (u = 0) when the proportion of infected individuals falls below the “opening threshold” which is reached when *I*(*t*) falls below 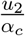. The social planner chooses the levels of mitigation 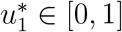 and 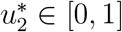 that minimize the overall fatality rate in the population *D*(*T*).

Results are in Table 1. As in the previous section, the fatality-minimizing levels of mitigation are below 100%. In all cases, acute mitigation reduces the proportion *I*(*t*) of infected individuals in the population, then mild mitigation ensures that this proportion remains approximately constant at a low level until time *T*. In the first six cases in Table 1, the minimum for *D*(*T*) is reached for *u*_1_ ≈45% and *u*_2_ ≈15%. When *α_c_* is higher, the social feasibility constraint in equation (8) is weaker, i.e., mitigation is more sustainable. In the last case in Table 1, a sufficiently weak social feasibility constraint (*α_c_* = 1000) allows a “suppression” strategy to be successful. This is achieved by strong acute mitigation (*u*_1_ ≈ 93%) followed by long-lived mild mitigation (*u*_2_ ∽ 53%), see the right panel in Figure 4. Even in this case, not switching to mild mitigation would result in the opening threshold being reached at some point, i.e., strong acute mitigation is not sustainable.

Indeed, in any case, setting the level of mild mitigation *u*_2_ too high would reduce the proportion *I*(*t*) of infected individuals too quickly, so that it would fall below the opening threshold prior to time T. As a result, mild mitigation would be lifted while the proportion of susceptible individuals, and therefore the effective reproduction number *R*, are still high, which would lead to a spike in infections. This would substantially increase the fatality rate, until acute mitigation is once again activated when the acute mitigation threshold is reached.

**Table 1:**
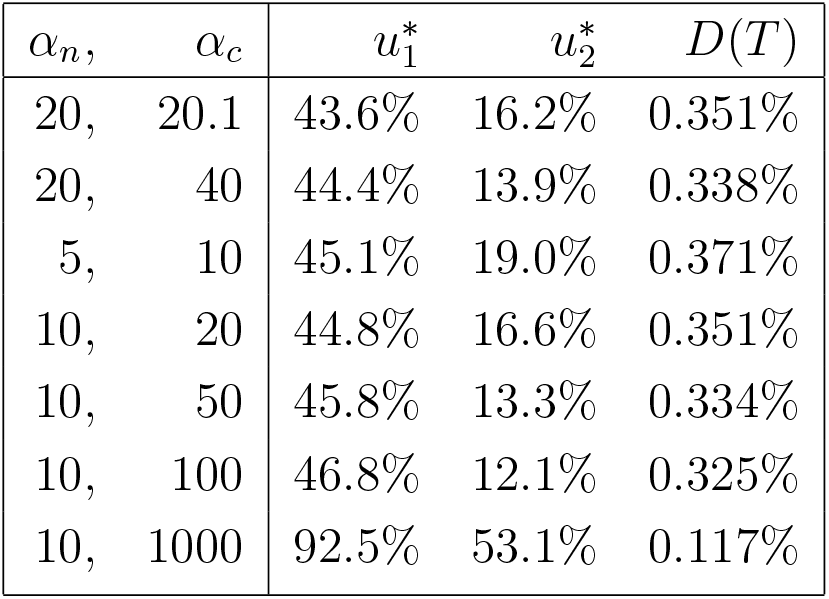
*Fatality-minimizing mitigation levels for several social feasibility parameters*

| $\alpha_n,$ | $\alpha_c$ | $u_1^*$ | $u_2^*$ | $D(T)$ |
| --- | --- | --- | --- | --- |
| 20, | 20.1 | 43.6% | 16.2% | 0.351% |
| 20, | 40 | 44.4% | 13.9% | 0.338% |
| 5, | 10 | 45.1% | 19.0% | 0.371% |
| 10, | 20 | 44.8% | 16.6% | 0.351% |
| 10, | 50 | 45.8% | 13.3% | 0.334% |
| 10, | 100 | 46.8% | 12.1% | 0.325% |
| 10, | 1000 | 92.5% | 53.1% | 0.117% |

**Figure 4:**
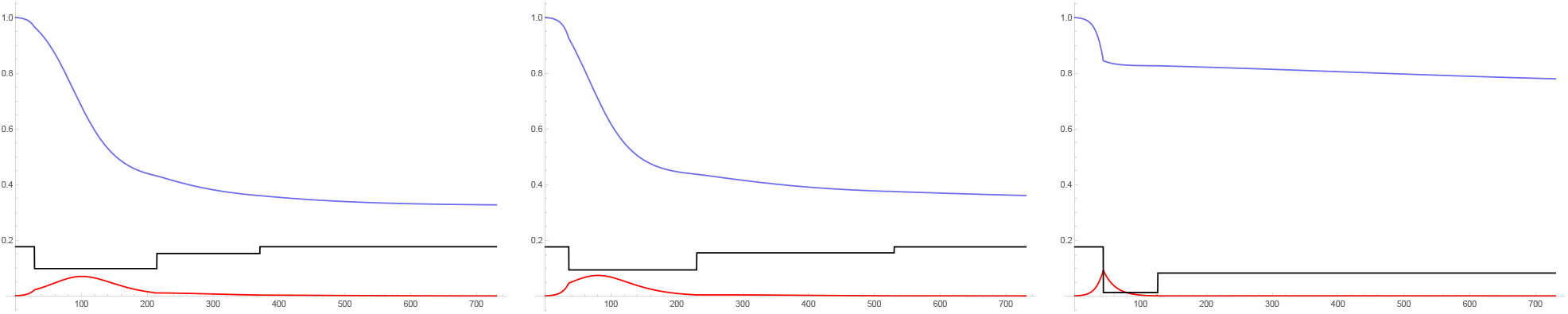
*I*(*t*) (red), *S*(*t*) (blue), and *b*(*t*) (black), with fatality-minimizing mitigation levels from Table 1. On the left, *α_n_* = 20 and *α_c_* = 40. In the middle, *α_n_* = 10 and *α_c_* = 100. On the right, *α_n_* = 10 and *α_c_* = 1000.

### 3.5 Three mitigation levels

This section considers policies with three mitigation levels, i.e., *u* can take four values: 0, *u*_1_, *u*_2_, and *u*_3_, with 0 ≤*u*_3_ < *u*_2_ < *u*_1_ ≤ 1. Acute mitigation starts (*u* = *u*_1_) when *I*(*t*) rises above 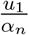 (see equation (7)), mild mitigation starts (*u* = *u*_2_) when continuing with *u* = *u*_1_ would violate social feasibility in equation (8), i.e., when *I*(*t*) falls below 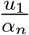, milder mitigation starts (*u* = *u*_3_) when continuing with *u* = *u*_2_ would violate social feasibility in equation (8), i.e., when *I*(*t*) falls below 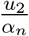, and mitigation ends (*u* = 0) when *I*(*t*) falls below 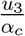. The social planner chooses the levels of mitigation 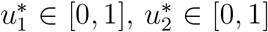, and 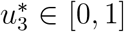 that minimize the overall fatality rate in the population *D*(*T*).

Results are in Table 2. Again, we find non-monotonic relations between mitigation and the fatality rate. Only with a very high social tolerance for mitigation policies (*α_c_* = 1000) is the level of acute mitigation 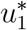 close to 100%. Even then, such strong mitigation is temporary (see the right panel in Figure 5), and it would have resulted in a higher fatality rate if it had not been followed in a timely manner by mild mitigation.

**Table 2:** Fatality-minimizing mitigation levels for several social feasibility parameters

| $\alpha_n,$ | $\alpha_c$ | $u_1^*$ | $u_2^*$ | $u_3^*$ | $D(T)$ |
| --- | --- | --- | --- | --- | --- |
| 20, | 20.1 | 45.6% | 22.7% | 9.6% | 0.345% |
| 20, | 40 | 46.5% | 19.8% | 7.7% | 0.332% |
| 5, | 10 | 48.5% | 27.1% | 12.3% | 0.364% |
| 10, | 20 | 47.1% | 23.2% | 9.8% | 0.345% |
| 10, | 50 | 48.6% | 19.7% | 7.6% | 0.329% |
| 10, | 100 | 57.9% | 33.2% | 17.6% | 0.308% |
| 10, | 1000 | 99.9% | 55.8% | 53.3% | 0.104% |

**Figure 5:**
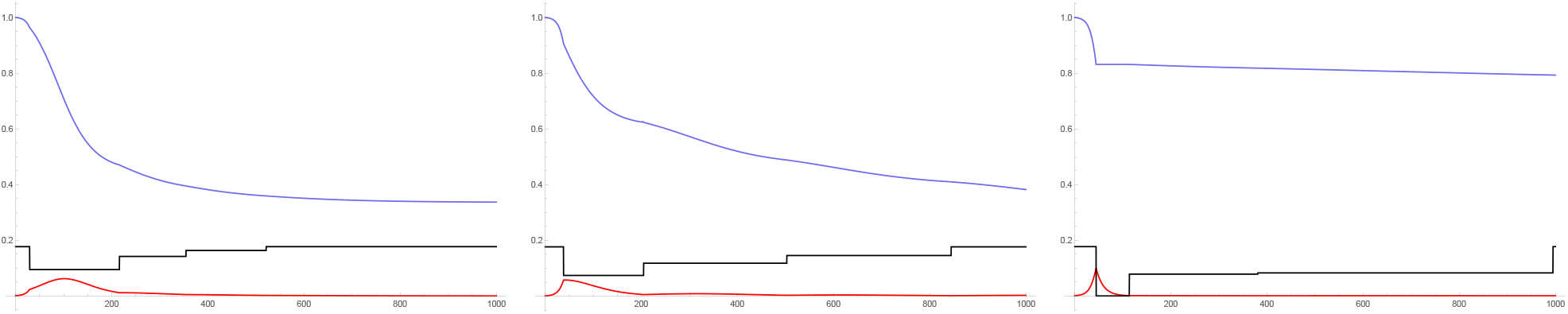
*I*(*t*) (red), *S*(*t*) (blue), and *b*(*t*) (black), with fatality-minimizing mitigation levels from Table 2. On the left, *α_n_* = 20 and *α_c_* = 40. In the middle, *α_n_* = 10 and *α_c_* = 100. On the right, *α_n_* = 10 and *α_c_* = 1000.

## 4 Discussion

### 4.1 Robustness

Our main results are independent of the nature of mitigation measures, the mitigation cost, and the postulated fatality rate for infected individuals. First, the analysis does not specify which mitigation measures are taken to reduce the effective contact rate: we focus on the level of mitigation rather than its specific nature. Second, the results are independent of the 10 mitigation cost, which is not considered. The analysis shows that, even with a zero mitigation cost, an excessive reduction in the effective contact rate could ultimately increase the overall fatality rate. Third, the main results are independent of the postulated fatality rate for infected individuals, *δ*. A higher *δ* would increase deaths proportionally in all cases (see equation (6)), so that it would not change the ordering of various cases by fatality rate.

The rest of this section studies the robustness of the results to two important assumptions.

First, results depend on assumptions made in the SIR model, most notably the R_0_ of the infectious disease. Figure 6 below is a robustness test in which we assume *R*_0_ = 4 and *R*_0_ = 5 (instead of *R*_0_ = 2.65 in the rest of the analysis), which results in *b* = 0.26 and *b* = 0.33 (instead of b = 0.18), respectively. As shown in Figure 6, an increase in R_0_ leads to higher fatality-minimizing mitigation levels, respectively 52.8% and 58.8% (instead of 39.9% with b = 0.18). Thus, the results change quantitatively but not qualitatively: the overall fatality rate is still non-monotonic in the mitigation level.

**Figure 6:**
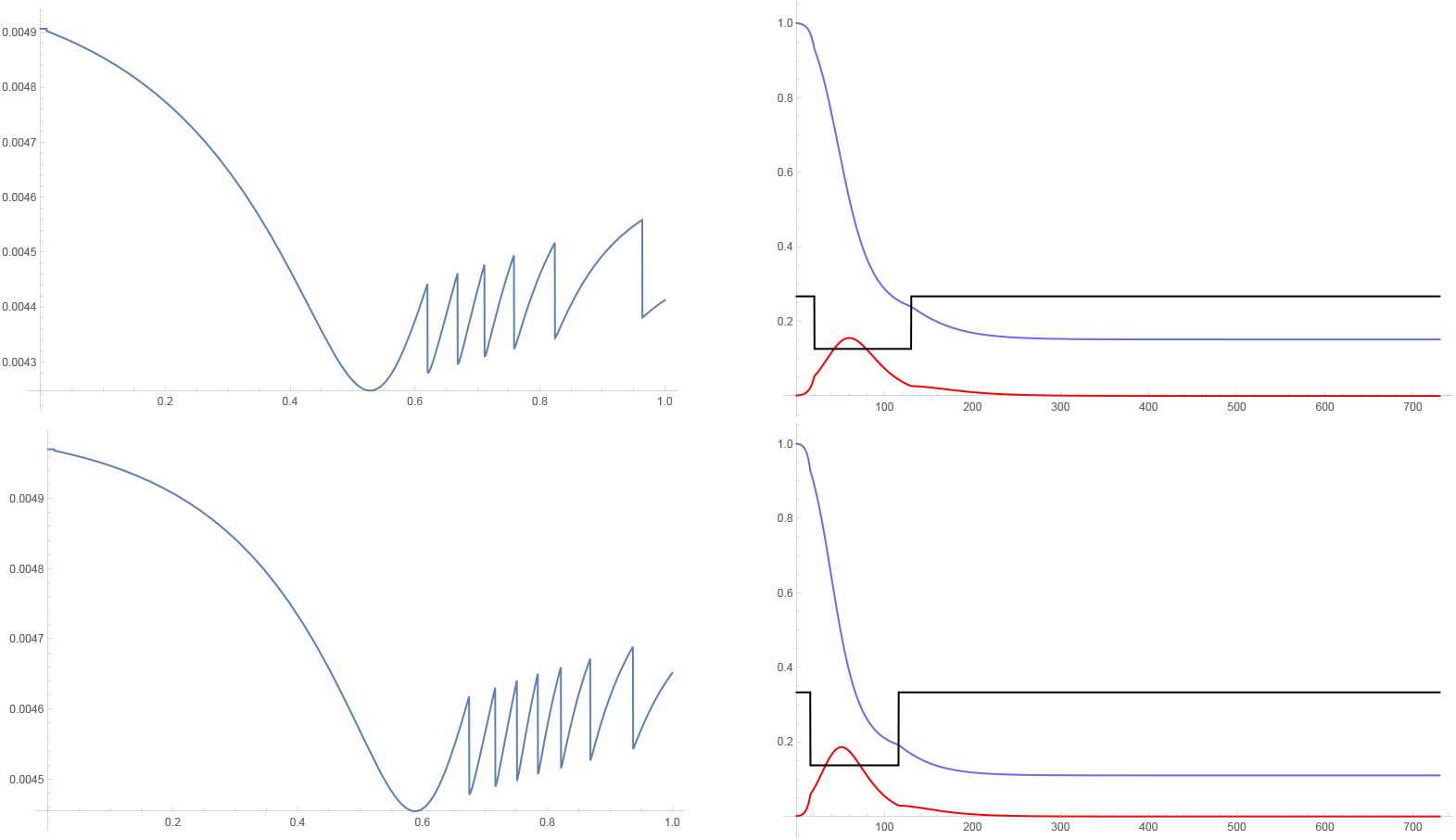
On the left, *D*(*T*) as a function of *u*_1_ for *u*_1_ ∈ [0,1] and *T* = 1, 000. On the right, *I*(*t*) (red), *S*(*t*) (blue), and *b*(*t*) (black) for the level 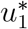 of *u*_1_ that minimizes the overall fatality rate. On both rows, *α_n_* = 10 and *α_c_* = 20. On the first row, *R*_0_ = 4, which gives 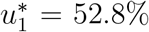, and *D*(*T*) = 0.425%. On the second row, *R*_0_ = 5, which gives 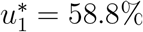, and *D*(*T*) = 0.445%.

Second, we address the robustness of results to the postulated exogenous ending time of the pandemic, time *T*. To take into consideration the fact that time *T* is unknown and hard to predict in advance, we now assume that *T* is uniformly distributed on 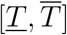. We are looking for the mitigation level *u*_1_ that minimizes the expected fatality rate in the population given this distribution. Formally, we consider the following function of *u*_1_:

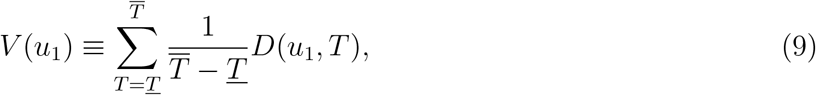

where the fatality rate in the population at time *T* is denoted as *D*(*u*_1_, *T*) to make clear that it depends on u_1_. In Figure 7, we plot *V*(*u*_1_) as a function of *u*^1^ ∈ [0, 1]. The fatality-minimizing mitigation level is very close to previous estimates when T is in-between 1 and 3 years 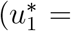, while it is higher when *T* could be only six months away at the beginning of the epidemic 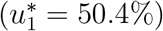. In any case, the fatality rate remains non-monotonic in *u*_1_.

**Figure 7.**
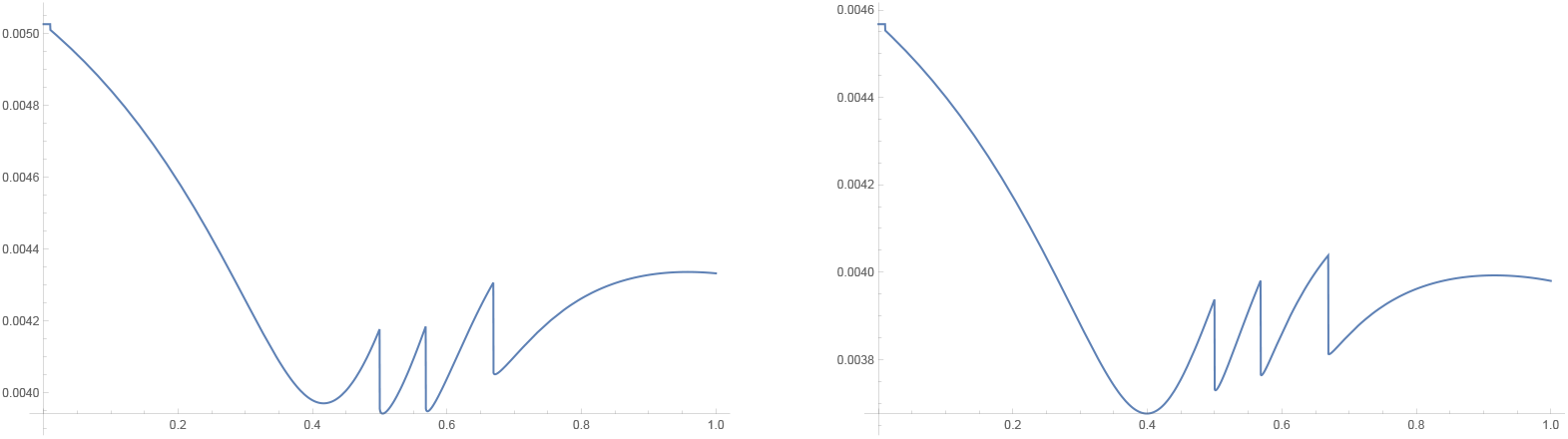
*V*(*u*_1_) as a function of *u*_1_ for *u*_1_ ∈ [0, 1] in the model with *α_n_* = 10, *α_c_* = 20 and one mitigation level *u*_1_. On the left, *T* ∽ *𝒰*[180, 730], and the minimum is reached at 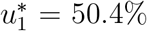, where the expected value of *D*(*T*) is 0.358%. On the right, *T* ∽ *𝒰*[365,1095], and the minimum is reached at 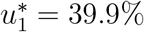, where the expected value of *D*(*T*) is 0.368%.

### 4.2 Inefficient policies

For policy design and evaluation purposes, it is instructive to compare the fatality-minimizing mitigation policies described above with alternative mitigation policies.

On the left panel of Figure 8, the postulated mitigation policy involves excessive mitigation, which results in a substantially higher overall fatality rate. By excessively slowing the spread of the pandemic, strong mitigation (*u*_1_ = 65%) brings down the proportion of infected individuals to a level such that the opening threshold is reached. Even though the proportion of infected individuals is low at this point, the proportion of susceptible individuals is still quite high because of the preceding strong mitigation. Thus, relaxing mitigation results in additional spikes in infections which trigger additional periods of mitigation (“stop and go” mitigation or “rolling lockdowns”).

On the right panel of Figure 8, the mitigation policy which is fatality-minimizing for *α_n_* = 10 and *α_c_* = 1000 is here inefficiently adopted when *α_n_* = 10 and *α_c_* = 100. Mitigation is too strong given the lower social feasibility. This results in a second spike in infections, a second mitigation period, and a relatively high overall fatality rate.

**Figure 8:**
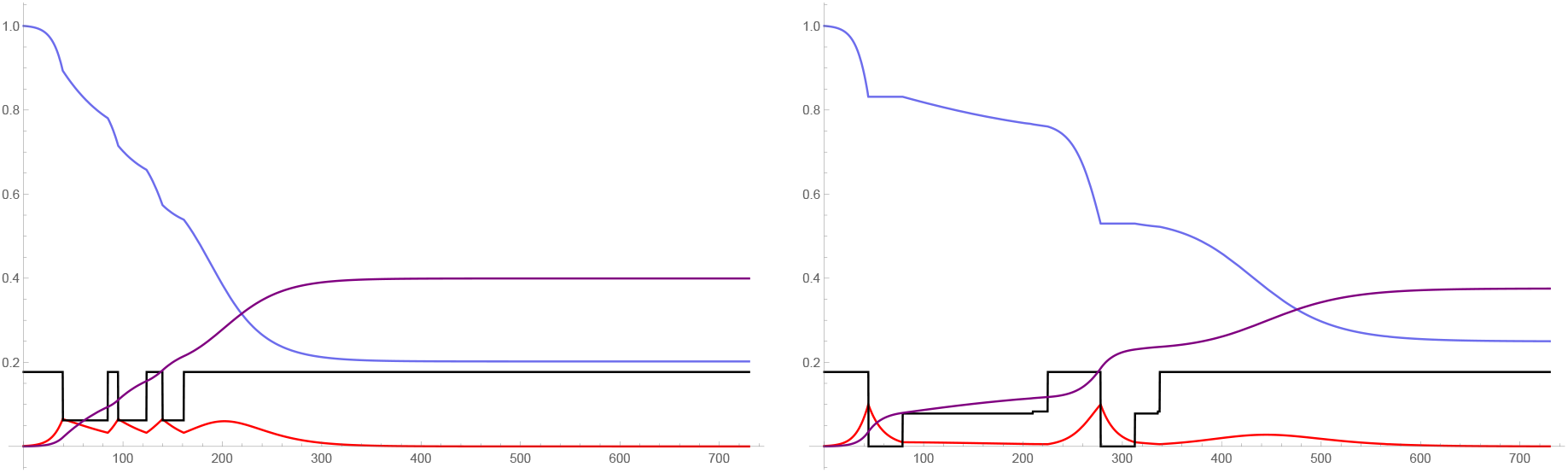
*I*(*t*) (red), *S*(*t*) (blue), *b*(*t*) (black), and 100 × *D*(*t*) (purple), for *T* = 1, 000. On the left, *α_n_* = 20 and *α_c_* = 40; *u_1_* is set at 65.0%, which gives *D*(*T*) = 0.399%. On the right, *α_n_* = 10 and *α_c_* = 100; *u*_1_ = 99.9%, *u*_2_ = 55.8%, and *u*_3_ = 53.3%, which gives *D*(*T*) = 0.376%.

## 5 Conclusion

Cricitism of mitigation policies taken in response to the COVID-19 pandemic have mostly focused on their economic cost. This paper highlights another aspect. If social acceptance of mitigation policies and individual compliance are only ensured when the infection rate is high enough, then excessive mitigation can backfire by excessively decreasing the infection rate and consequently undermining the social feasibility of future mitigation. That is, even without any explicit mitigation cost, increasing the level of mitigation beyond a certain point can still be detrimental.

This new effect, presented here in a simple and transparent SIR model, could be incorporated into more complex models that take into account additional factors as well as local conditions. The social feasibility constraints could be extended to take a more general form, for example with time-varying social feasibility parameters, a nonlinear specification, or with additional state variables besides the rate of infection in the population. The set of feasible policies could also be extended beyond threshold policies. Finally,it is an open question whether the limited scope and duration of mitigation policies are in practice primarily dictated by economic costs, by constraints on the social feasibility of mitigation policies, or by a combination of these two considerations.

## Data Availability

There is no data.

## Supplementary Online Appendix

### Not for publication

#### Social feasibility and COVID-19

In France, it is only after the number of deaths from COVID-19 increased substantially that citizens started socially distancing, even though the situation in precursor countries such as China and Italy had been widely reported in the news. In France, “the first day of the national lockdown [March 17] did not go according to plan. In some big cities (notably Paris), large crowds could be seen wandering in the streets. Shoppers bustled through the streets; few wore masks or kept the one-metre distance.” Only after the situation “dramatically worsened” did compliance increase.^10^ This was short-lived: a few weeks after the peak in COVID-19 cases which in France was on March 31, compliance had noticeably decreased, especially in relatively spared rural areas, and almost two-thirds of French people were ready to resume normal life.^11^

In California, which was relatively spared from COVID-19 infections and deaths, compliance with instructions to socially distance was low: “shortly after [Governor] Newsom issued his directive, crowds packed beaches stretching along California’s coastline (…) Parks were jammed with visitors and farmers markets bustled with shoppers.”^12^ By contrast, New York state suffered a high number of COVID-19 infections and fatalities. Its population was largely compliant with mitigation measures, especially in the epicenter of the outbreak, New York city.^13^ Yet, only a few weeks after mitigation measures started, and after the number of COVID-19 cases and deaths started declining, the state started facing growing social pressure to lift mitigation measures: “local leaders [are] anxious to reopen schools and businesses. [New York state Governor Andrew] Cuomo said he knows local officials are under pressure to act.”^14^

In several countries, including the US, the UK, France, Spain, and Italy, compliance with lockdown orders diminished as lockdowns stayed in place and COVID-19 related cases and deaths declined, a phenomenon referred to as “lockdown fatigue”.^15^ In the US, where COVID-19 cases rapidly increased at the end of March and have been on a downward trend since the beginning of April, cell phone data shows that the level of social distancing at the end of April reverted to its March 20th level in many states.^16^ In the UK, a few weeks after the peak on April 8 but while the lockdown was still in place, numbers of vehicles on the road increased and visits to parks “almost returned to normal levels”.^17^

#### Social feasibility parameters

We discuss the assumption *α_c_* > *α_n_*. First, this condition is realistic: all else equal, social feasibility is usually a greater concern when a new policy is put in place than when it is continued. Second, this condition is without loss of generality. Indeed, with *α_c_* ≤ *α_n_*, mitigation could not start when equation (7) is binding: if it did, when *I*(*t*) reaches the point when mitigation can start with a given positive value for *u*(*t*), it is also at a level such that mitigation must end because of equation (8), a contradiction. Mitigation u can only start and last when *I*(*t*) is above both 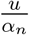 and 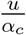 (see equation (7) and equation(8)), so that there is no loss of generality in setting *α_c_* ≥ *α_n_*. Moreover, with *α_c_* = *α_n_:* either mitigation starts when equation (7) is binding, but then it would end at the same time as it would start, i.e., there is no mitigation; or mitigation could start when equation (7) is not binding, but this is equivalent to having *α_c_* > *α_n_*.

#### Exogenous thresholds

We study what happens when the mitigation threshold and the opening threshold are independent of the mitigation level u_1_, for example because they are set in advance or because the social planner has a “window of opportunity” to engage in mitigation. In Figure 9, we simply use the thresholds that would correspond to *α_c_* = 10 and *α_n_* = 20 with u_1_ = 40%, and hold these thresholds constant. That is, the mitigation level only has an effect on the intensity of mitigation, but not on the mitigation period(s). The simulation depicted in Figure 9 shows that the overall fatality rate is still non-monotonic in the level of mitigation *u*_1_, and is minimized at 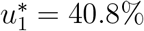.

**Figure 9:**
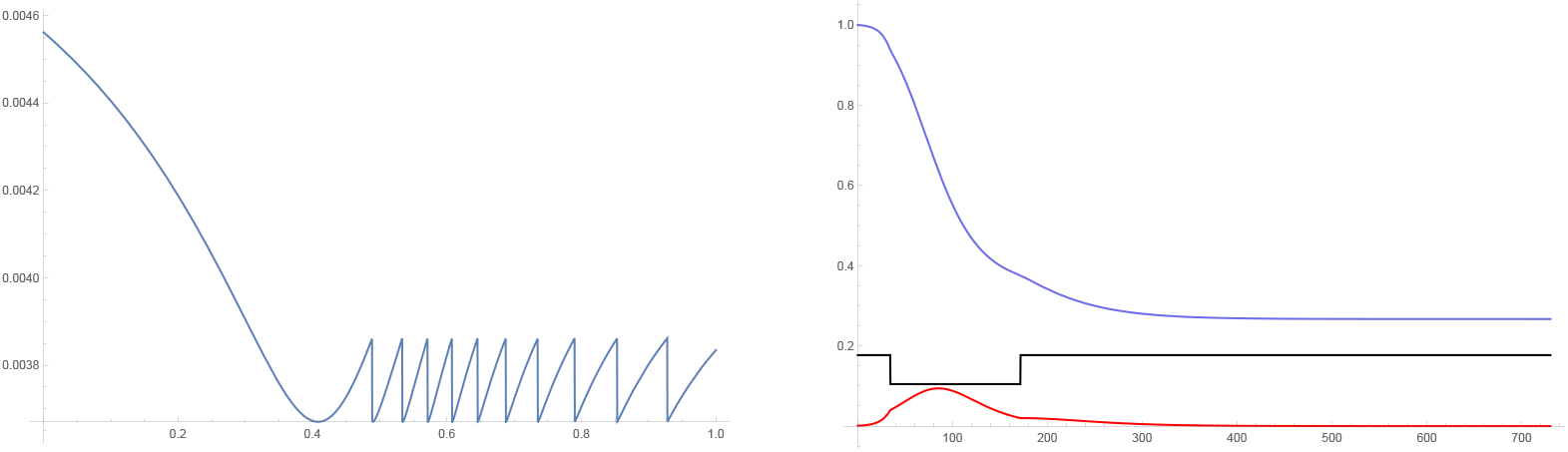
The mitigation threshold is 4% and the opening threshold is 2%. On the left, *D*(*T*) as a function of *u*_1_ for *u*_1_ ∈ [0, 1] and *T* = 1, 000. On the right, *I*(*t*) (red), S(t) (blue), and *b*(*t*) (black) for 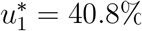, which gives *D*(*T*) = 0.367%.

#### Two mitigation levels: additional analysis

**Figure 10:**
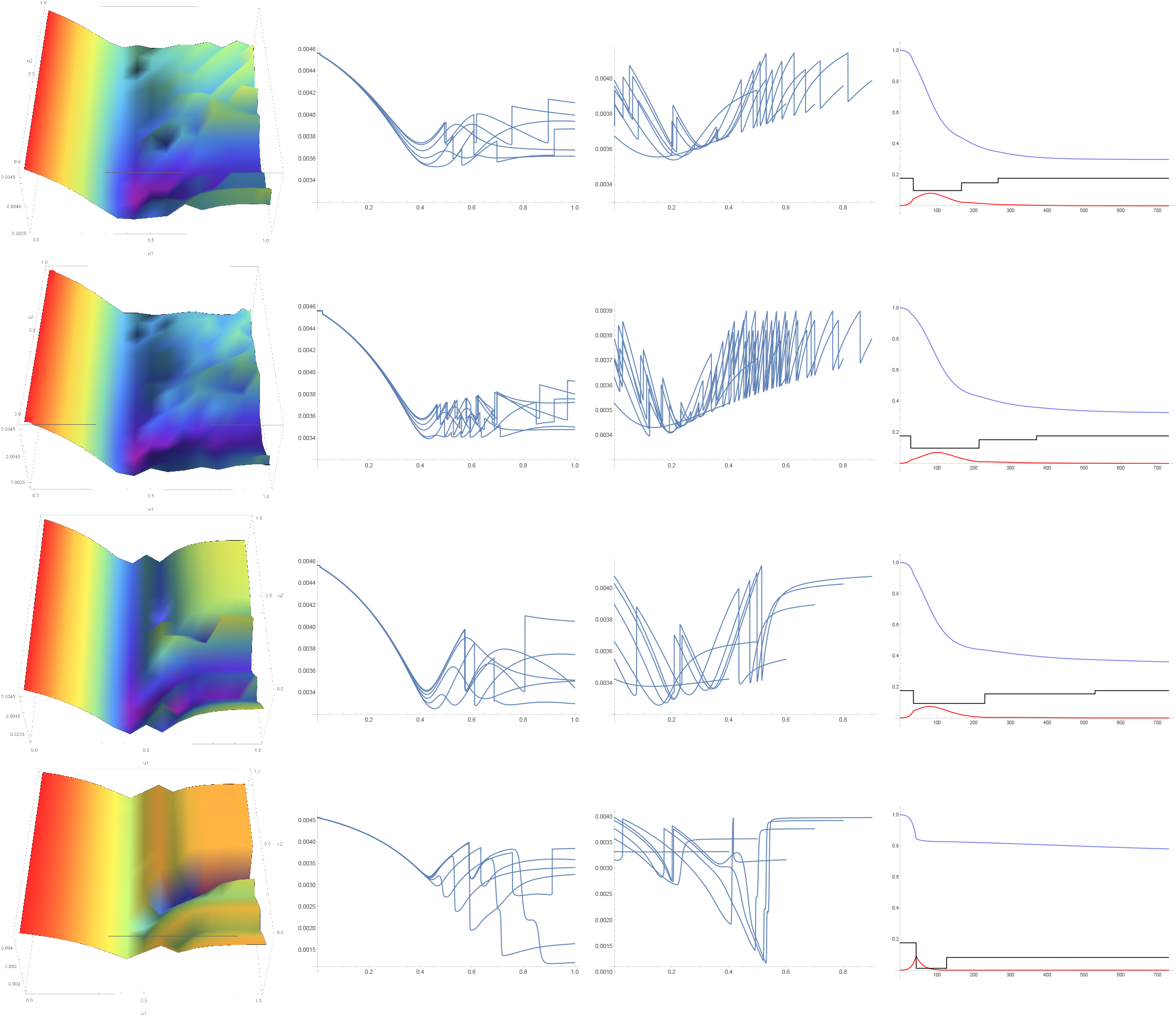
On the left, *D*(*T*) as a function of *u*_1_ ∈ [0,1] (horizontal axis) and *u*_2_ ∈ [0,1] (vertical axis), and *T* = 1, 000. On middle left, *D*(*T*) as a function of *u*_1_ ∈ [0,1] for *u*_2_ ∈ {0.1, 0.2, 0.3, 0.4, 0.5, 0.53} and *T* = 1, 000. On middle right, *D*(*T*) as a function of *u*_2_ ∈ [0,1] for ui ∈ {0.4, 0.5, 0.6,0.7, 0.8, 0.9}, and *T* = 1, 000. On the right, *I*(*t*) (red), *S*(*t*) (blue), and *b*(*t*) (black) for the levels 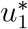 and 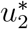 of *u*_1_ and *u*_2_ that minimize the overall fatality rate – see Table 1. On the first row, *α_n_* = 10 and *α_c_* = 20. On the second row, *α_n_* = 20 and *α_c_* = 40. On the third row, *α_n_* = 10 and *α_c_* = 100. On the fourth row, *α_n_ =* 10 and *α_c_ =* 1000.

#### End time *T*: additional analysis

In Figure 11 below, we calculate the value 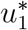 of *u*_1_ that minimizes the overall fatality rate in the population *D*(*t*) in 1,000 simulations where *T* is uniformly distributed on [180,1460] and is known ex-ante. We find that 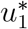 is close to 40% in around 80% of cases, and the highest value of 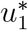 is 59.1%.

**Figure 11:**
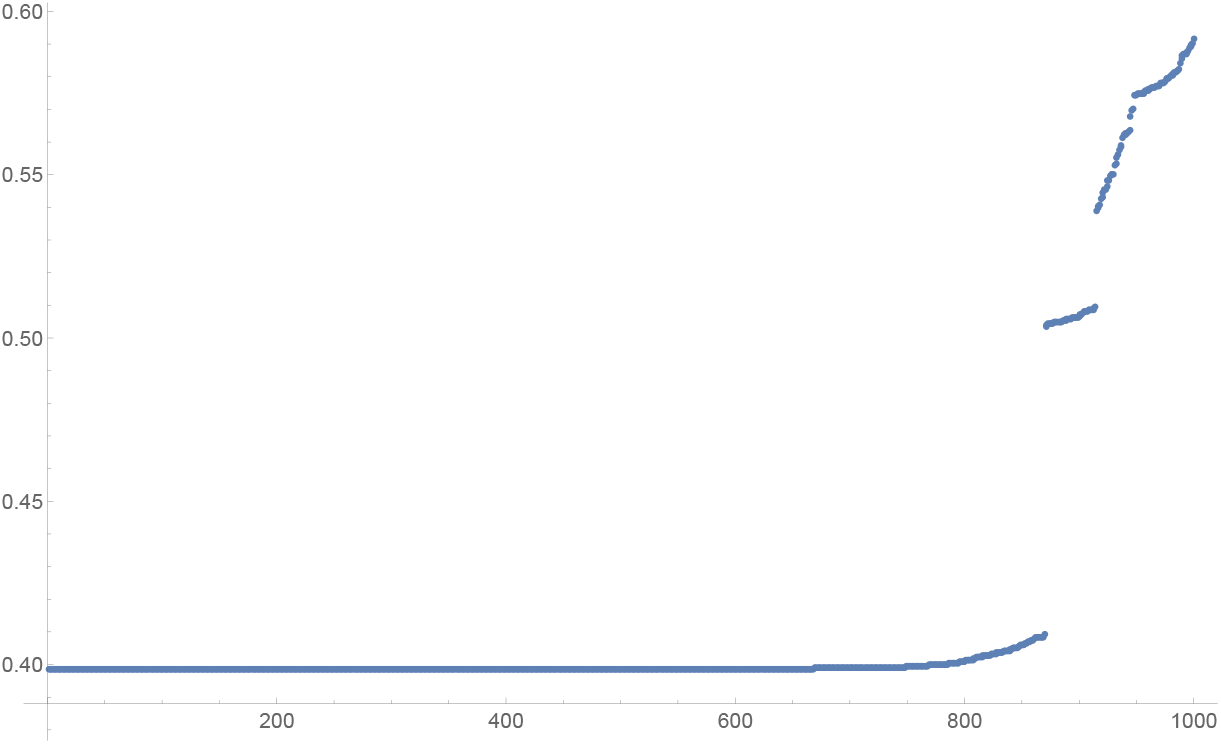
Below are the ordered values of *u*_1_ that minimize *D*(*T*) in the model with *α_n_* = 10, *α_c_* = 20 and one mitigation level *u*_1_, with 1,000 simulations involving different values of *T* when *T* is uniformly distributed on [180, 1460] and the realized value of *T* is known at the time of setting *u_1_*.

## Footnotes

People do not fully internalize the effect of their decision on the spread of the virus, which suggests a role for mandatory lockdowns to address externalities (Eichenbaum, Rebelo, and Trabandt (2020)).

This is what China did to beat coronavirus. Experts say America couldn’t handle it, USA Today April 1 2020.

This is related to the notion of “reactive social distancing”, which postulates that individuals spontaneously alter their behavior in response to increased infection risk (Reluga (2010), Perrings et al. (2014), Yu et al. (2017),Baker (2020), Toxvaerd (2020)).

“When will it be over?”: An introduction to viral reproduction numbers, R0 and Re, April 14 2020.

Antibody tests support what’s been obvious: Covid-19 is much more lethal than the flu, Washington Post April 28 2020.

This is what China did to beat coronavirus. Experts say America couldn’t handle it, USA Today April 1 2020.

China’s aggressive measures have slowed the coronavirus. They may not work in other countries, Science magazine March 2 2020.

Politicians can’t hide behind scientists forever - even in a pandemic, LSE blog, May 6 2020.

According to equation (2), the proportion of infected individuals remains constant or declining over time (*R* ≤ 1) if and only if: 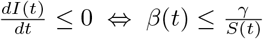 Using *β*(*t*) = *b*(1 − *u*) from equation (5), the level of mitigation u such that *I*(*t*) is constant or declining over time is: 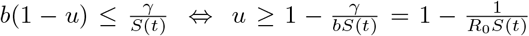, where the last equality uses *b* = *γR*_0_ in a SIR model. Thus, for *u* = 0 to be consistent with a constant or declining proportion of infected individuals over time, we must have 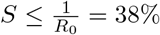with *R*_0_ = 2.65.

Source: The French are finally observing lockdown advice – but is it too late? The Guardian March 19 2020.

Source: A l’approche du déconfinement, il y a de plus en plus de monde dehors, Le Figaro May 2 2020.

Source: Public Health Officials To Newsom: Lockdown Won’t Work Without Enforcement, Kaiser Health News, March 26 2020.

Source: Desolate New York: eerie photos of a ghost metropolis, The Guardian, April 4 2020.

Source: Cuomo on reopening: Can’t be stupid, more people will die if we’re not smart, Syracuse.com

Source: London pedestrian traffic jumps as UK coronavirus lockdown fatigue sets in, City AM, April 28 2020., French public feels lied to as lockdown fatigue grows, Financial Times April 16 2020., Lockdown fatigue hits asEurope enforces coronavirus restrictions, Financial Times April 1 2020.

Source: Is quarantine fatigue here? Americans are leaving their homes more and more, cell data shows, USA Today April 27 2020.

Coronavirus: Eight ways the lockdown has changed the UK, BBC News, May 1 2020.

## Notes

### Competing Interest Statement

The authors have declared no competing interest.

### Funding Statement

No funding required.

